# Impacts of Global School Feeding Programmes on Children’s Health and Wellbeing Outcomes: A Scoping Review

**DOI:** 10.1101/2024.09.03.24312981

**Authors:** Amy Locke, Michaela James, Hope Jones, Rachel Davies, Francesca Williams, Sinead Brophy

## Abstract

**Background:** School feeding programmes (SFP) are one of the world’s most extensive interventions to alleviate child poverty, food insecurity and malnutrition. Schools are ideal settings to promote healthy eating habits early in life since children have access to at least one main meal per day at school. However, there is a lack of clear evidence for the effectiveness of school feeding programmes on child health outcomes. Additionally, there is considerable debate on whether feeding programmes should be universal or targeted with countries taking different approaches. This review will therefore explore global research on school feeding programmes and consider different health outcomes through targeted or universal policies.

**Methods:** We conducted a search across four electronic databases. These studies investigated the impacts of school feeding programmes on children’s health outcomes. 207 papers were identified with 76 studies subjected to full text screening.

**Results:** A total of 42 papers were included in the final review. All papers were published within the past fifteen years (2009-2024) and included thirteen countries in total. SFP were associated with child weight being more in the healthy range. Targeted SFP were negatively associated with stigma.

**Conclusions:** Universal SFP were effective at improving children’s health outcomes such as healthy weight, improved behaviour and social support. Overall, both targeted and universal SFP positively impact children’s health outcomes and address health disparities.

**STRENGTHS AND LIMITATIONS OF THIS STUDY:**

- The review mapped both universal and targeted provisions, providing a broad overview of the existing literature.
- The review included an appraisal of the methodological quality of the included studies. Only English studies were included.
- Only studies published in English were included, potentially excluding relevant research in other languages.
- The study did not perform a formal assessment of publication bias, which could impact the interpretation of the results.

## Introduction

School feeding programmes (SFP) are one of the world’s most extensive interventions to alleviate child poverty, food insecurity and malnutrition [1]. In 2022, 418 million children worldwide received a free or subsidised school meal as part of a SFP [2]. In recent years, growing concerns have arisen regarding the nutritional health of children [3]. With increased sugar intake, increases in childhood obesity and socio-economic factors affecting millions of families worldwide, there is emphasis on the need for interventions to reduce negative health outcomes [4]. Schools present an opportunity to promote healthy eating habits among children and can serve as a preventative measure to negative health implications [5].

From a public health perspective, schools are ideal settings to promote healthy eating behaviours early in life since children have access to at least one main meal per day at school in most schools [6]. There is a large body of literature dedicated to exploring the impacts of SFP on educational outcomes, noting positive outcomes, specifically in relation to key stage attainment and attendance [7, 8]. However, literature relating to the impacts of SFP on children’s health outcomes are limited, and even less research considers SFP as a preventative tool for public health. Research by Chaudhary et al [9] demonstrated that initiatives targeting food and nutrition within schools could enhance dietary habits, promote healthy eating, and impact body measurements positively. Noting that interventions with a focus on meal provision, healthy eating promotion and food literacy are most effective in improving children’s health outcomes.

It is well documented that good nutrition plays a vital role in maintaining good health throughout the life course, preventing malnutrition in all forms, and decreasing the risk of non-communicable diseases, such as cardiovascular disease, diabetes, and some cancers [10]. Increases in processed foods and changes in lifestyles have led to a shift in dietary patterns, with individuals opting for cheaper food options and convenience foods [11]. With the rise in poor dietary habits, there is a negative impact on health outcomes. This has become a public health concern, specifically in relation to rising childhood obesity. Figures from 2022 indicate childhood obesity has reached an all-time high with an estimated 390 million children and adolescents aged 5–19 years being overweight or living with obesity. Of this, 37 million children under age five were classified as overweight [12, 13]. To date, no country is on track to curb the obesity crisis [14]. Moreover, in 2019, over 161 million children under five years of age were reported as having a nutritional deficiency globally from undernourishment [15]. Nutritional deficiencies have been attributed to childhood wasting, stunting, infections, as well as cognitive and behavioural disorders [16, 17, 18].

Health disparities due to poor nutritional intake is largely seen in poor economic groups [19], and often arise in early childhood, resulting in on-going implications into adulthood [20]. According to the World Health Organisation (WHO) [5], childhood and adolescence is a critical period for promoting nutritional health and reducing the risk of negative health implications and calls for policy reform to SFP to address diet quality in children are becoming widespread [21].

Despite the large potential health benefits of SFP, they have proven difficult to evaluate given the diversity in implementation policies across nations. For example, Europe alone has a range of SFP policies from universal to targeted approaches [22]. Universal systems refer to a provision open to all students, regardless of socio-economic status [23]. This system is well established in countries such as Sweden, where all children in both primary and secondary education (ages 5 – 16) have access to at least one free school meal (FSM) a day and is regarded as a symbol of national welfare [24]. However, there is also diversity among policies relating to universal systems, whereby countries offer universal school meals to children in particular year groups. For instance, Latvia offers free meals to children in grades one to four (ages 7 –9), and Lithuania provides FSM from preschool to first grade (up to age 7) [22].

Targeted programmes on the other hand, target children from lower socio-economic backgrounds, usually using a means-tested eligibility system which is based on parental income [25]. This approach is implemented in Poland, Slovenia, and parts of the United Kingdom [26]. This approach, whilst beneficial to some families, has its own limitations, e.g. a threshold cut off means families slightly above the eligibility criteria may still be living in poverty but are able to access FSM and the stigma associated with being eligible may lead to a lack of uptake [23].

There is considerable debate among government policymakers as to whether SFP should be targeted (benefit eligibility depends on family income) or universal (benefits are provided to all students with no eligibility criteria) [27, 28]. However, given the notable gap in health-related literature, it is difficult to establish which system, if any, is most effective at addressing health disparities.

To our knowledge, no study has systematically synthesised information relating to the effectiveness of SFP on children’s health outcomes. Therefore, this review aims to address the following questions; 1. To what extent does the existing literature examine the impacts of school feeding programmes on the physical, emotional, psychological, and social health of school children globally and 2. What challenges, including factors influencing uptake, are reported in the literature regarding the implementation of free school meal provision and 3. Does the effectiveness of SFP on children’s health outcomes vary depending on the SFP policy (targeted or universal).

For the purpose of this scoping review, children’s health and wellbeing will be categorised as physical health, emotional and psychological health, and social health.

## Methods

### Review Design

We conducted a scoping review of both qualitative and quantitative research to establish evidence of the effectiveness of school feeding programmes in improving children’s health outcomes globally. This scoping review was conducted in accordance with the Preferred Reporting Items for Systematic Reviews and Meta-Analysis (PRISMA) guidelines [29]. The protocol for the review was registered on the Open Science Framework (OSF) [30]. A preliminary search of MEDLINE, the Cochrane Database of Systematic Reviews was conducted, and no current or underway systematic reviews or scoping reviews on the topic were identified. The study findings were summarised through a narrative synthesis. Researcher (AL) extracted study data from full text papers. Each extraction was reviewed by a second reviewer. Any conflicts were presented to and resolved through group discussion.

### Search Strategy

Four electronic databases – Medline, PubMed, Web of Science and Google Scholar (including Science Direct, Web of Science, ProQuest Central, EBSCO, CINAHL) – were searched to identify relevant research papers in English. Studies published from 2009 were included, with most papers from 2020 to 2023. The literature search was conducted by lead author (AL) in December 2023 with screening taking place in January and February 2024. Additionally, the reference lists of all included sources were also screened for additional studies. Reports and case studies were also sourced and included in the final screening. Search terms included: Universal Free School Meals OR School Feeding Programmes OR Free School Meals and Children’s Health OR Free School Meals and Children’s Wellbeing OR School Feeding Programmes OR Challenges with School Feeding Programmes OR School Meal Provision. Additionally, each string was searched using the phrase ‘Impacts of.’

### Eligibility

#### Inclusion Criteria

Studies exploring school feeding programmes were included if they met the following outcomes: (1) Children’s physical health – Illness, physical activity, and/or diet dietary intake. (2) Children’s emotional and psychological health– emotion regulation, resilience, self-esteem, and behaviour. (3) Children’s social health – social engagement/participation, building and maintaining relationships. (4) Studies published within the past fifteen years to ensure research is up-to-date and relevant. (5) Primary and secondary school aged children (ages 5 to 16). (6) Studies investigating the implementation of universal feeding programmes and its potential challenges. Both experimental and quasi-experimental study designs including randomised controlled trials, non-randomised controlled trials, before and after studies and interrupted time-series studies. Descriptive observational study designs including case series, individual case reports and descriptive cross-sectional studies were also included. Qualitative studies were also included that focused on children’s self-assessed health and wellbeing and perceptions of school feeding programmes.

#### Exclusion Criteria

Studies were excluded if they met the following criteria: (1) Children below or above school age (below age 5 or above 18). (2) Studies with a focus on SFP and academic outcomes – performance, attainment, or attendance without a health element. (3) Studies that only looked at parental funded meals/packed lunches. (4) Studies not in English and, (5) Studies published prior to 2009. Additionally, any studies that did not focus on the impacts of school meal provisions on children’s health, unless considering factors influencing uptake, were excluded.

### Evidence Selection

Selected papers were uploaded to Covidence systematic review software [31], by the lead author (AL). After removing 30 duplicates, the remaining titles and abstracts (177) were screened against the selection criteria by authors (AL, MJ, HJ, RD, SB). Remaining studies undertook full-text screening by reviewers (AL, MJ, HJ, RD, FW). Any conflicts during the review process were resolved through group discussion. One paper was removed during data extraction as it became apparent that it was an abstract for a conference presentation and no full text was available.

### Quality Assessment

Methodological quality of included studies was assessed using the Mixed Methods Appraisal Tool (MMAT) [32]. The MMAT is a quality appraisal tool developed to evaluate the methodological quality of empirical studies. The MMAT was first published in 2009. Since then, it has been validated in several studies testing its interrater reliability, usability, and content validity [33]. The tool can appraise five different categories of study designs: qualitative, randomised controlled trial, non-randomised, quantitative descriptive and mixed methods studies. As our review includes numerous study designs, this tool was deemed as appropriate. Each section is assessed based on five criteria for methodological quality, offering three responses: “Yes” (indicating the criterion is satisfied), “No” (indicating the criterion is not satisfied), and “Can’t tell” (suggesting insufficient information to assess). Studies are categorised as low quality if they receive a “Yes” response for two or less questions, moderate quality for 3 questions, and high quality for four or more questions.

Four papers included in the review were impact reports and opinion pieces, and did not fit under an appraisal category within the MMAT, therefore the modified version of the Joanna Briggs Institute (JBI) critical appraisal checklist for text and opinion was used to quality assess these papers [34]. Appraisal is subjected to the same response criteria as above; “Yes” (indicating the criterion is satisfied), “No” (indicating the criterion is not satisfied), and “Can’t tell” (suggesting insufficient information to assess). No papers were excluded based on the results of the quality assessment. Quality assessment was conducted by one reviewer (AL) and checked by a second reviewer(s).

### Data Extraction and Analysis

Relevant information from selected studies was extracted using a template (this is provided in the Supplementary Information) developed by lead author (AL). The data collection template included information on; author(s), year of publication, Country, type of meal provision, participants, study design, and key findings, gaps/limitations identified and recommendations for future research. The development of the template was guided by the JBI manual for evidence synthesis [35] and piloted on Covidence using two papers.

Data analysis consisted of a narrative synthesis due to heterogeneity of both study designs and outcomes among included studies, making statistical meta-analysis impractical. Analysis and synthesis were conducted following recommendations from the Guidance of the Conduct of Narrative Synthesis in Systematic Reviews [36]. Specifically, a grouping strategy was utilised to arrange findings into similar theme groups.

## Results

### Overview of studies

The literature search yielded a total of 207 titles, and these were imported to Covidence for title and abstract screening. After duplicates were removed (n= 30), 177 titles and abstracts remained for screening. Of these, 101 were excluded for not meeting the inclusion criteria, i.e., wrong outcomes measured. A further 34 studies did not meet the eligibility criteria and were excluded at full text screening phase, with a majority vote for wrong outcomes studied. A total of 42 papers were included in the final review. Figure 1 illustrates the screening process.

**Fig 1.**
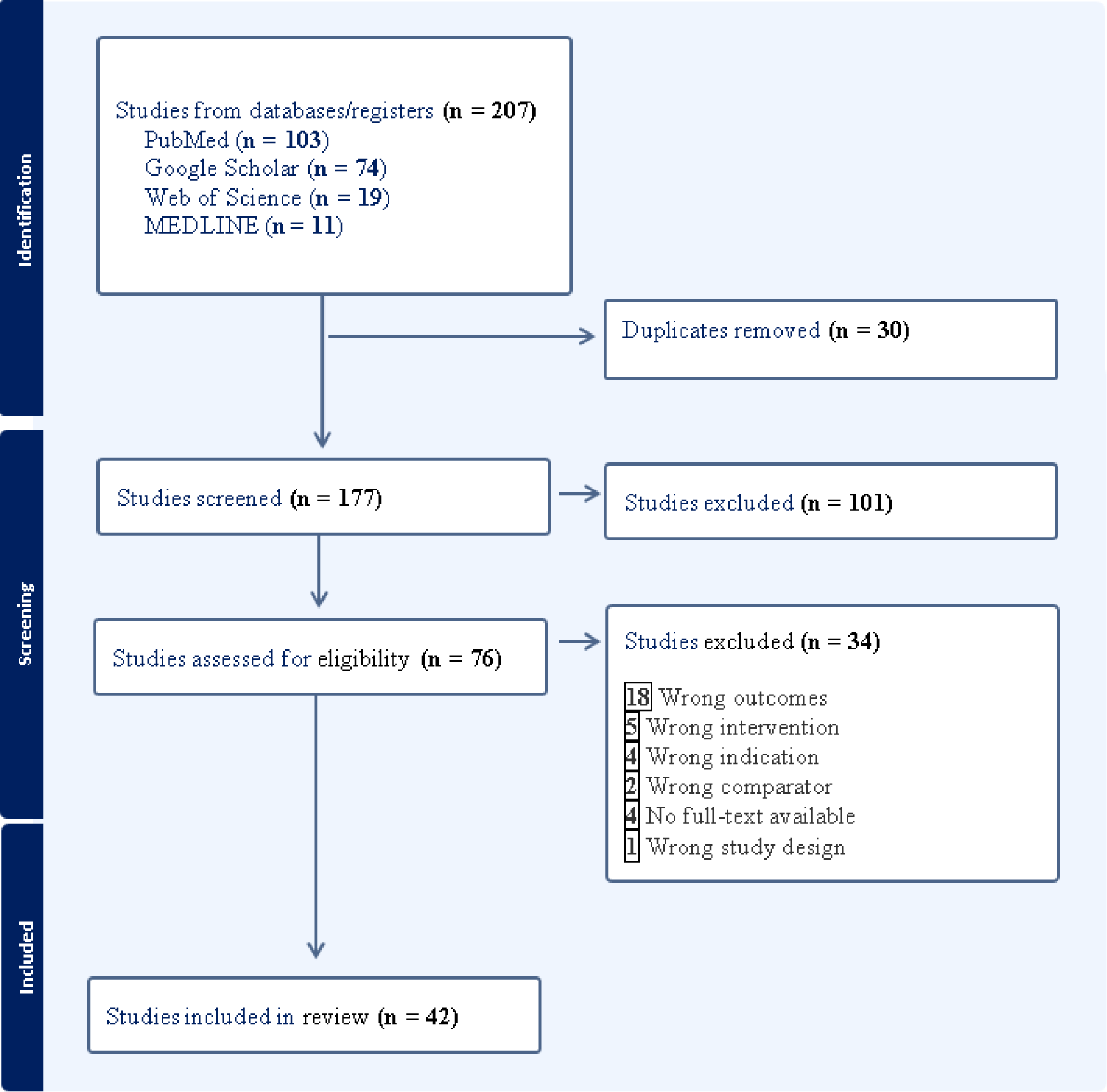
PRISMA Diagram.

#### Quality of Included Papers

Using the modified version of the Joanna Briggs Institute (JBI) critical appraisal checklist for text and opinion [34], three papers were categorised as high, and the other was categorised as moderate. The three papers categorised as high [24, 69, 72], used a variety of methodologies or a strong evidence base to support their findings/opinion. The paper rated moderate [70] included less peer reviewed evidence to support their findings but methodological quality was satisfactory.

#### Characteristics of Included Studies

Characteristics of the included studies are summarised in table 3. 11 studies were cross-sectional [37–47]. Five studies had a difference-in-difference design [20, 49–51]. Three were non-randomised control trials [6, 52, 53]. Four were quantitative descriptive [54–57]. Four were mixed methods [58–61]. Six were qualitative [62, 64–68]. Six were reports and/or opinion pieces [22, 24, 70–72]. One was a rapid narrative review [73] and two were case studies [63, 74].

**Table 1:** MMAT Quality Assessment results.

| **Study** | **Overall Score** |
| --- | --- |
| Altindag et al. [57] | Moderate |
| Batista et al. [37] | Moderate |
| Bethmann and Cho [48] | High |
| Cardoso et al. [62] | High |
| Jessiman et al [65] | Low |
| Chambers et al. [63] | High |
| Chambers et al. [74] | High |
| Colombo et al. [38] | High |
| Davis et al. [57] | Low |
| Evans [39] | High |
| Garton et al. [73] | High |
| Goel et al. [55] | Moderate |
| Goodchild et al. [40] | Moderate |
| Guio [22] | High |
| Hecht [59] | High |
| Holford and Rabe [49] | Low |
| Holford and Rabe [50] | High |
| Horta et al. [41] | High |
| Illøkken et al. [64] | Low |
| James [42] | Low |
| Carlisle et al. [58] | High |
| Long et al. [43] | High |
| Lundborg et al. [20] | High |
| Mauer et al. [66] | High |
| McKelvie-Sebileau1 et al. [67] | High |
| Meier et al. [54] | Low |
| Neervoort et al. [52] | Low |
| Parnham et al. [51] | High |
| Parnham et al. [44] | Low |
| Rahim et al. [71] | High |
| Sahota et al. [68] | Moderate |
| Spence et al. [45] | Moderate |
| Taylor et al. [60] | Low |
| Vik et al. [6] | High |
| Vik et al. [53] | High |
| Yamaguchi et al. [61] | High |
| Yang et al. [46] | High |
| Zailani et al. [47] | High |

**Table 2:**
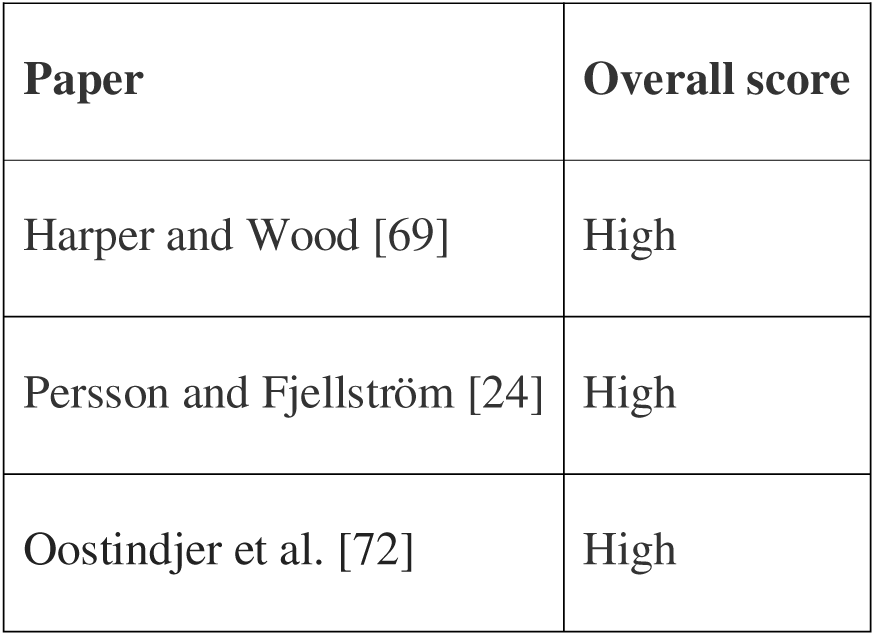
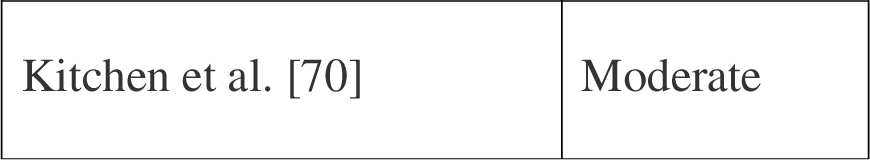
JBI critical appraisal checklist for text and opinion results.

**Table 3:** Characteristics of the included studies.

| Author | Country | Provision<br>Type | Participant<br>School year | Design |
| --- | --- | --- | --- | --- |
| Altindag et al.<br>[57] | South Korea | Universal | All | Empirical analysis |
| Batista et al. [37] | Brazil | Targeted | Aged 4-14 | Cross-Sectional |
| Bethmann and<br>Cho [48] | South Korea | Universal | Aged 9-16 | Difference-in-Difference |
| Cardoso et al.<br>[62] | Portugal | Targeted | Secondary | Qualitative |
| Jessiman et al<br>[65] | London | Universal | Secondary | Mixed Methods |
| Chambers et al.<br>[63] | Scotland | Universal | N/A | Qualitative Case<br>Study |
| Chambers et al.<br>[74] | Scotland | Targeted | Secondary | Case Study |
| Colombo et al.<br>[38] | Sweden | Universal | Secondary | Cross-sectional |
| Davis et al. [56] | Georgia-USA | Universal | All | Descriptive |
| Evans [39] | England | Other | Primary | Cross-sectional -<br>Observational |
| Garton et al. [73] | New Zealand | Universal | Not specified | Rapid Narrative<br>Review |
| Goel et al. [55] | USA - | Other | Primary | Observational |
|  | Virginia |  |  |  |
| Goodchild et al.<br>[40] | England | Universal | Primary | Cross-sectional -<br>Descriptive |
| Guio [22] | Europe | Other | N/A | Policy<br>Recommendation |
| Harper and Wood<br>[69] | UK Wide | Targeted | Primary | Report |
| Hecht [59] | USA | Universal | All | Mixed methods |
| Holford and Rabe<br>[49] | England | Universal | Primary | Difference-in-Difference |
| Holford and Rabe<br>[50] | England | Universal | Primary | Difference-in-Difference |
| Horta et al. [41] | Brazil | Targeted | Primary | Cross-sectional –<br>Analytical |
| Illøkken et al.<br>[64] | Norway | Targeted | Secondary | Qualitative |
| James [42] | England | Targeted | Secondary | Cross-sectional -<br>Descriptive |
| Carlisle et al. [58] | London | Universal | Not specified | Qualitative |
| Kitchen, et al. | England | Universal | All | Impact report |
| [70] |  |  |  |  |
| Long et al. [43] | USA | Universal | All | Cross-sectional |
| Lundborg et al.<br>[20] | Sweden | Universal | Birth to 50+ | Difference-in-Difference |
| Mauer et al. [66] | Norway | Targeted | Secondary | Qualitative |
| McKelvie-Sebileau et al.<br>[67] | New Zealand | Universal | Secondary | Qualitative |
| Meier et al. [54] | USA | Targeted | N/A | Quantitative<br>Exploratory |
| Neervoort et al.<br>[52] | Kenya | Universal | Primary | Non-randomised<br>Control Trial |
| Oostindjer et al.<br>[72] | Cross-National | Other | N/A | Discussion/Essay |
| Parnham et al.<br>[51] | England and<br>Scotland | Universal | Primary | Difference-in-Difference |
| Parnham et al.<br>[44] | UK | Targeted | All | Cross-sectional |
| Persson &<br>Fjellström [25] | Sweden | Universal | All | Article piece |
| Rahim et al. [71] | England | Universal & Targeted | Primary | Government Report |
| Sahota et al. [68] | England | Targeted | All | Qualitative |
| Spence et al. [45] | England | Universal (Infant) | Primary | Cross-sectional |
| Taylor et al. [60] | Vermont, USA | Universal | Staff | Mixed methods |
| Vik et al. [6] | Norway | Targeted | Secondary | Non-randomised Control Trial |
| Vik et al. [53] | Norway | Universal | Secondary | Non-randomised Control Trial |
| Yamaguchi et al. [61] | Japan | Universal | Primary | Mixed methods |
| Yang et al. [46] | UK | Targeted | All | Cross-sectional |
| Zailani et al. [47] | Nigeria | Targeted | Primary | Cross-sectional |

Twenty-three papers were based on universal school feeding programmes [20, 24, 38, 40, 43, 45, 48, 49, 50, 51, 52, 53, 56, 57, 59, 60, 61, 63, 65, 66, 67, 70, 73]. Fifteen had targeted SFP systems [6, 37, 41, 42, 44, 46, 47, 54, 62, 64, 66, 68, 69, 72, 74]. Four papers were categorised as ‘other’. Papers were categorised as other if they looked at breakfast [55], compared both universal and targeted systems [71], compared packed lunch to school meals [39] or were not specified [22].

Nine papers included children in all school ages (primary and secondary) [24, 43, 44, 46, 56, 57, 59, 68, 70], Twelve papers included children just in primary schools [39, 40, 41, 45, 49, 50, 51, 52, 55, 61, 69, 71], Ten papers included children just in secondary schools [6, 38, 42, 53, 62, 64, 65, 66, 67, 74]. Two papers had an overlap of different age groups (children aged 4-14 [37] and children aged 9-16 [48]). One study looked at the impacts of SFP across the life course (from birth to 50+) [20]. One paper included school staff [60]. Five papers did not explicitly state age groups [22, 54, 63, 65, 72].

Seventeen papers were from UK (England n=11 [39, 40, 42, 45, 49, 50, 58, 65, 68, 70, 71], Scotland n= 2 [63, 74], England and Scotland n=1 [51], UK wide n=3 [44, 46, 69]. Six papers were from the USA [43, 54, 55, 56, 59, 60], three papers were from Sweden [20, 24, 38], four from Norway [6, 53, 64, 65], one from Portugal [62] one was Europe wide [22]. Two were from New Zealand [67, 73], two from Brazil [37, 41], two from South Korea [47, 48], two from Africa (Kenya [52] and Nigeria [47], one from Japan [71] and one was cross national [72]. Figure 2 demonstrates the countries of the included papers.

**Fig 2.**
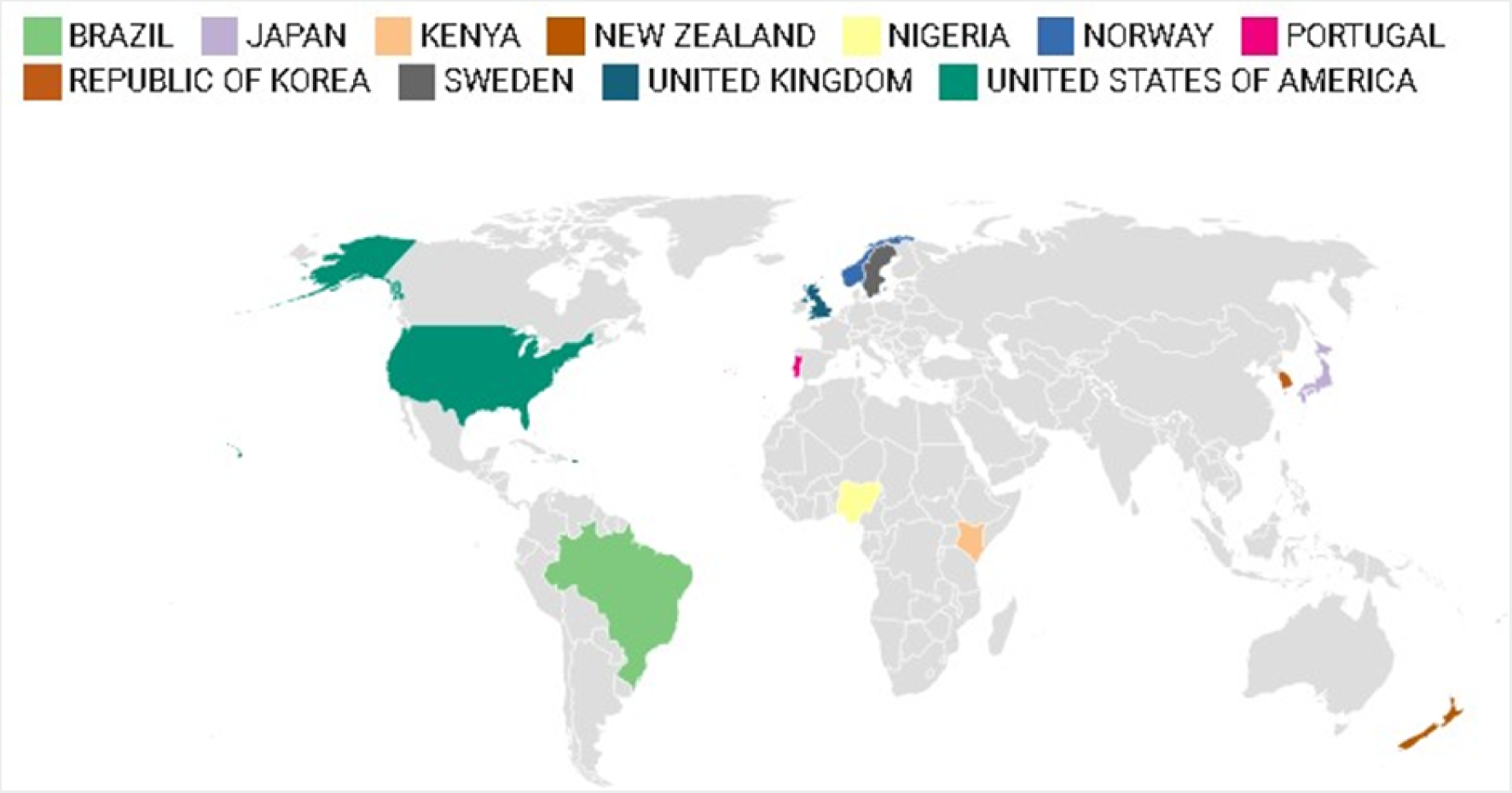
Map showing countries included in review.

Overall studies that investigated universal feeding programmes appear to have the most significant positive outcomes on children’s health, specifically in relation to behaviour, health, and socio-economic outcomes. Whilst targeted systems adequately address food insecurity and child hunger, children eligible for targeted provision had a higher probability of experiencing poor mental health and stigma. Findings are discussed in more detail below.

#### Physical Health: BMI and Body Weight Outcomes

Five papers investigated the impacts of SFP on children’s BMI and weight outcomes. Two with targeted provision [6, 37] and three with a universal system [48, 50, 56]. Studies had contradicting results.

Batista et al, [37], investigated the prevention of overweight children in Brazil through targeted SFP. The study which utilised a cross-sectional design and included 7,017 participants from 21 schools found that being overweight was prevalent in 30.6% of children and being underweight was present in 1.9%. This study highlighted that a large proportion of the foods served were ultra-processed, recommending a re-evaluation of food standards within schools. Researchers noted an inability to observe extracurricular activities relating to food consumption and physical activity which may also influence children being overweight.

Bethman and Cho [48], investigated the impacts of reintroducing universal free school meals (UFSM) in South Korea after the policy had been abolished a year prior. Findings indicated the removal of the policy had adverse effects on children’s BMI outcomes. The reintroduction of the policy on the other hand reversed these impacts and saw more children with healthier weights. However, the sample size of the study is not explicitly stated and may not represent the intended population.

Holford and Rabe [50], suggested that longer exposure to a universal infant SFP positively influenced children’s weight outcomes. The study included 153,522 primary school children aged between four and seven and used a difference-indifference analysis to measure children’s weight outcomes over time. Findings concluded that children are more likely to be a healthy weight after exposure to the provision over the course of the first year of school. Though participants were very young, it is recommended that participants are tracked over a longer time period to see if this is a lasting effect.

A study by Vik et al, [6] conducted in a secondary school in Norway, investigated changes in children’s dietary habits, BMI and waist circumference modulated by socio economic status. The study conducted a non-randomised control trial consisting of two groups, non-exposure to SFP and exposure to SFP under a targeted system. Findings suggested free school meals (FSM) increased the intake of healthy foods, particularly among children with lower socio-economic status, leading to an increase in the Healthy Food score of the intervention group between baseline and follow-up. However, children in the intervention group had a significant increase in BMI, though it is not explicitly stated if the children were underweighting prior to the trial. The study concluded that interventions to promote healthy eating are more effective in lower socio-economic groups if it is free or at reduced price.

Davis et al. [56] saw similar results when they examined the effects of a universal SFP in a deprivation area in the USA. Reporting that BMI increased in provision schools versus non provision schools. They further stated that children in provision schools had a higher BMI on average.

In conclusion, there is no evidence that SFP lower BMI, but children are more likely to be a healthy weight. Factors such as the prevalence of ultra-processed foods, socio-economic status, and policy reintroduction played significant roles in these outcomes. Longer exposure to SFP, particularly universal programs, was associated with healthier weight outcomes in younger children, though lasting effects require further study.

#### Nutritional Intake

Ten papers [24, 53, 51, 39, 45, 61, 52, 47, 38] explored the impacts of SFP on nutritional intake. Outcomes explored were diverse. An Article piece [24] claimed that Sweden was one of the countries with the highest vegetable intake, with a substantial intake at lunch. Sweden has the longest running universal SFP in the world and supplies food to all school aged children. The article notes that school meal provision in Sweden is seen as a universal welfare service and a part of public health work.

Vik et al. [53] used a non-randomised control trial including 164 secondary school children to investigate whether serving a free healthy school meal for one year resulted in a higher intake of fruit and vegetables and a lower intake of unhealthy snacks. Vik et al. [53] reported that a free healthy school meal for one year was associated with higher weekly intake of vegetables on sandwiches in the intervention group compared to the control group. However, it was not associated with a lower weekly intake of unhealthy snacks in the intervention group compared to the control group. It is important to note that the study had a small sample size and relied on self-reported data.

Parnham et al. [51] examined the impact of a universal infant SFP policy on dietary intake in primary school children in England and Scotland. The implementation of the policy led to more children participating in school meals. No impact was seen on fruit and vegetable intake but there was evidence that the policy lowered consumption of foods associated with packed lunches, such as crisps, and some nutrients, such as total fat and sodium.

A further study by Evans [39] also reported that school meals were a healthier option than packed lunches. The study utilised a cross-sectional design and included 2709 primary school children in England. Findings suggested children having a packed lunch consumed on average 11g more total sugars than those who had a school meal whereas, children having a school meal were more likely to consume different types of vegetables and drink water. Overall, children with a packed lunch consumed a lower-quality diet over the whole school day. The study did not look directly at school meal provision and so it is difficult to note whether those on provision would yield differing results.

Spence et al. [45] explored the effect of pre– and post-Universal Infant Free School Meals (UIFSM) and school on pupil’s dietary intakes using a cross-sectional research design. At lunchtime, there was a statistically significant decrease in students’ non-milk extrinsic sugars intake pre implementation and increases of intakes of cake’s pudding post implementation. However, only two schools were included, thus limiting the generalisability of results. It is recommended that schools consider healthier policies when implementing SFP.

Yamaguchi et al. [61] indicated that universal SFP were beneficial to individuals with low socio-economic backgrounds. Their study which used a mixed methods approach and included 719 primary school children in Japan, saw that children whose mothers were less educated had greater reliance on school lunch for their vegetable intake. Children with lower household income had more contribution from school lunch to their fruit intake. Household income was not explicitly considered in the study and mothers’ education was used to determine socio-economic status, so it is unclear whether families were from lower socio-economic backgrounds. Further studies are needed to conclude the correlation between vegetable intake and household income in Japanese children.

Four papers, Neervoort et al. [52], Zailani et al. [47], Colombo et al. [38] and Goel [55] considered the impacts of SFP on vitamin intake as well as nutritional deficiencies.

Neervoort et al. [52] conducted a non-randomised control trial including sixty-seven primary school children to investigate the impacts of a universal SFP on anaemia, stunting, wasting and malnutrition. Findings indicated the programme reduced anaemia and malnutrition, and improved child growth in the study group. Whilst improvements were seen, the study had several limitations. Such as a very small sample size and participants were from only one school. The study also lacked information on the content and nutritional value of provided meals, which could influence program effectiveness.

Zailania et al. [47] investigated portion sizes of targeted school meals as well as nutrient intake and found that the meals served through the SFP contributed at least 33% for energy, protein, iron, calcium, sodium, vitamin A, and zinc intake. However, there was no consumption of fruits, meat, poultry, and fish within the programme. The study did not consider the contribution of other food sources, including snacks and homemade meals which may account for nutritional values.

Colombo et al. [38] used a cross-sectional design to explore School Lunch Dietary intake under a universal SFP in Sweden. Results found that a quarter of the overall energy intake; between 22% and 30 % of selected nutrient intakes; almost half of vegetable intakes; roughly two-thirds of fish intakes; and around a third of red/processed meat intakes. These findings imply that school meals make an important contribution to children’s diets on weekdays. Researchers also reported school meals as more nutritious than meals consumed outside of school, though meals consumed outside of school meals were not measured.

Goel et al. [55] examined total sugar in free breakfasts served in elementary (primary) schools in Virginia, USA through an observational study. Findings suggest that meals offered might contribute to excessive overall sugar availability for children. Nutritional information for added sugars was not provided within the study, however, the findings may explain contradictions in BMI above. Goel et al. [55] states districts and policymakers should collaborate to implement more effective guidelines concerning sugar availability in breakfast items to optimise children’s dietary intake.

In summary, the evidence for improved nutritional intake is limited. However, school meals are an important contribution to diets during weekdays. Consuming school meals alleviated dietary discrepancies related to social inequalities.

#### Psychological and Emotional Health Outcomes

The impacts of SFP on children’s psychological and emotional outcomes were investigated in four papers [57, 60, 48, 46].

A randomised control trial by Altindag et al. [57] evaluated the effects of targeted school meals on students’ behaviour, bullying, and violence in schools. Behavioural incidents reduced by 35% since the implementation of a universal system in 2010 (particularly physical fights among students). It is believed that this reduction is the result of reduced stigma among students as socio-economic status cannot be identified via the system. Provision did not impact students’ nutritional intake but reduced the stigma of receiving free meals through a targeted system by mitigating the possibility of identifying peers’ socioeconomic statuses.

Taylor et al. [60] included 116 school staff in the USA to explore the impact of a universal SFP on school climate, behaviour, and attainment. Taking a mixed-methods approach, over 725 of school staff surveyed reported that serving universal school meals has improved social climate as well as reductions in student stress, family financial stress and school administrator stress. This is thought to be a result of income differences being less visible, and the school community feeling more inclusive. It is recommended that further research include interviews with students and families to gain a better understanding.

As previously mentioned Bethman and Cho [48] saw positive results when looking at children’s BMI. Similarly, the reintroduction of a universal policy also saw mental health improvements among students. Proclaiming that free school lunches help to improve health and benefit student welfare. However, mental health was assessed using a ‘crying without any reason’ measure. The measure used is not reported so validity and reliability cannot be determined. No other Mental Health outcomes were assessed.

A further paper by Yang et al. [46] assessed targeted school meals in the UK on children’s mental health outcomes. The study used a cross-sectional design and included 2166 primary and secondary children. Yang et al [46] reported that poor mental health was observed in food insecure children receiving provision but also food insecure children not receiving provision. Interestingly, food insecure (i.e., not having access to sufficient food) children receiving provision had a higher probability of poor mental health than those who were food insecure and not receiving provision. It is thought that this is because targeted approaches are accosted with stigma.

In conclusion, universal approaches were associated with improved mental health outcomes, such as reduced behavioural incidents in school and an improved social climate. This was not observed with targeted approaches.

#### Social Health Outcomes

Five papers [58, 65, 64, 66, 68] investigated the social impacts of SFP. The impacts of SFP are multifaceted with outcomes seen with social interactions, reduced stigma and narrowing inequalities. Findings are discussed further below.

A mixed method, quasi-experimental evaluation exploring the impact of universal provisions in two London secondary schools by Carlisle et al [58] saw improved social skills as children were eating with peers. Improved behaviours, less stigma as well as students eating more varied foods. These findings were as a result of paired student interviews, this may have caused socially desirable responses.

A further study by Jessiman et al. [65] which also included a sample of secondary school students in London also reported social health benefits, Students perceived feeling equal under a universal system whereas they reported feeling ‘weak’ under the targeted system.

Illøkken et al. [64] conducted a qualitative study in a Norwegian secondary school, involving 13 students and five teachers to investigate the effects of a targeted school feeding program on social inequalities. Participants viewed school meals as a social event where students made new friends and learned new skills. It was also reported that social equality among students increased. However, it is unclear whether this was the result of the feeding programme or school meals in general.

Another qualitative study conducted in a Norwegian secondary school by Mauer et al. [66], also reported positive social outcomes. Students expressed that social time while eating school meals was important to them. This study also noted that popularity of the food was also important for attracting students to school meals.

Sahota et al. [68] investigated factors influencing take-up of targeted school meals through focus groups with children in both primary and secondary schools in England. Utilising focus groups to gain children’s perspectives. Food choice, queuing, and the social aspects of lunch time, such as eating with friends was a major influence in uptake. Recognition of the importance of the social aspects of dining for pupils and facilitation of social interactions through the spatial (including flexible locations, e.g. outside) and temporal organisation of lunchtimes. It is reported that the schools involved in the study had a high level of targeted school meal entitlement which may have resulted in the normalisation of school meal uptake. It is recommended that social aspects of school meal provision be further investigated in schools with lower levels of eligibility to see if results differ.

#### Reducing Food Insecurity and Child Hunger

Three papers [73, 59, 41] looked at the impacts of SFP on food insecurity and child hunger. Most papers in this review touched on food insecurity, however, these three papers specifically looked at impacts of food insecurity outcome. A rapid narrative review by Garton et al. [73] concluded that SFP in New Zealand significantly reduced hunger and food insecurity in primary schools. Hecht [59] suggested that access to free meals through a universal provision in a high deprivation area in the USA, reduced food insecurity among children as well as decreased child hunger. Lastly, a cross-sectional analysis by Horta [40], investigated impacts of SFP on Vulnerability risk. Findings saw positive impacts from consuming school meals on children’s diets, particularly among children living in high/very high social vulnerability risk areas.

#### Factors Influencing Uptake

Ten papers explicitly investigated SFP participation and factors associated with non-take-up among students. Two studies gained perspectives through qualitative methods [67, 62]. A paper by McKelvie-Sebileau et al. [67], involving universal provision, found that lack of knowledge of the programme and loss of agency over meal choices were major drivers of non-take-up but, positive influences such as participants’ food security, better nutritional knowledge and improved wellbeing were also perceived. Cardosa et al. [62] who evaluated a targeted approach in Portugal noted that quality of food was a concern when participating in school meals. Children who were not entitled to free or discounted meals reported eating at school less often than children with free or discounted lunches.

Three reports, Kitchen et al. [70], Rahim et al. [71] and Harper and Wood [69], all reported increased take-up under a universal policy, for both previously non eligible children and eligible children. It is suggested that improvements were seen as a result of reduced stigma and familiarising parents with school meals. Participants believed the pilot increased the range of food that pupils would eat, built their social skills at mealtimes and, for some pupils, resulted in health benefits associated with having a balanced meal, such as more energy, concentration and alertness and improved complexion. take-up was higher among primary school children than secondary school children. However, Rahim et al. [71] in particular, found schools experienced difficulty predicting the appropriate quantities of food required for each menu option, which meant that some meals either ran out early or were wasted.

A study by Holford and Rabe [49] also found that take-up of school meals in England by non-eligible children rose from a consistent 30-35% in the eight years preceding the policy to approximately 85% in the universal period.

Additionally, two case studies [74, 63], one exploring a targeted system and the other a universal system saw similar results. Under the targeted provision [74] stigma, food quality and social aspects played a vital role in take-up. Results specifically noted that, being separated from friends decreased take-up. Implementation of a universal system [63] however, yielded more positive results such as, families who were previously above the eligibility threshold now being able to access provision. It was also documented that school-provided meals were of a higher nutritional quality than those from home, exposing children to a greater variety of foods and establishing healthier eating habits.

Finally, a cross-national comparison by Oostindjer [72] indicated that removing popular foods such as meat from school menus significantly reduced take-up of school meals. This was particularly evident in Finland where meat and fish were removed from the school menu on some days reducing participation rate, consequently producing up to 60% plate waste. Please note that this paper did not explicitly mention SFP but instead, school menu choices and take-up therefore, more research is needed on food choices under SFP to evaluate if there is an effect on take-up.

#### Wider Impacts

The remaining seven papers did not fit into a thematic category and will therefore be summarised individually below.

A UK wide study by Parnham et al. [44] exploring access to targeted SFP, reported that receiving an FSM was associated with increased odds of recently using a food bank [75] but not reporting feeling hungry. In the month after the COVID-19 lockdown, 49% of eligible children did not receive any form of FSM. In this study the sample size was small, and this may lead to chance findings or in ability to detect differences.

A policy recommendation report by Guio [22], explored evidence of the short and longer-term benefits of school meal provisions in EU countries. The report found that health outcomes are dependent on the programme take-up and quality of food. Additionally, while eating formally, pupils learn to be sociable and develop interaction skills. There are some findings on health outcomes associated with school meal provision, but these are more difficult to establish. School-based food and nutrition interventions were able to improve dietary behaviour, healthy eating, and anthropometry, but the design of the intervention affects the magnitude of the effect.

James [42], used a cross-sectional design to investigate stigma associated with targeted provision and how this influences the peer effect. The study included 21,000 secondary school students in England. According to James [42], the presence of stigma dampens the peer effect and so students may not be benefiting from provision as much as possible. However, information about the provision has the opposite impact on the peer effect. These findings suggest that information is a more important part of the peer effect for those living in areas of greater deprivation and stigma is more important for those in the least deprived regions.

Long et al. [43] assessed the impacts of a universal system on meal cost and food quality in 508 US schools. This study finds that participation in the provision was associated with lower per-meal full cost with no differences in dietary quality. This indicates that in the US, universal SFP can provide nutritious meals to more students without a financial disadvantage. Meier et al. [54] conducted a quantitative exploratory study including 576 parents. Completed in the USA, the study aimed to gain parents overall perceptions of the targeted school meals program. This study found that parents of enrolled children were more likely to report positive perceptions of the school meals’ program. Whereas parents of children not receiving the program were less likely to perceive the school meals program in a positive light. Two limitations were noted that may have influenced results. Firstly, Parents of children not receiving provision made up much of the sample and second, parents volunteered to participate in this survey, therefore participation bias could have resulted from those who are more involved or have strong opinions about school lunch.

A longitudinal study, utilising a difference-in-difference design by Lundborg et al. [20] evaluated the long-term impacts of the universal program on children’s economic, educational, and health outcomes throughout the life course. The study involved 1,529,760 participants from birth to age 50+. The study noted several findings, for instance, children exposed to the program during their entire primary school period have 3% higher lifetime income. This effect was greater for pupils that were exposed at earlier ages and for pupils from poor households, suggesting that the program reduced socioeconomic inequalities in adulthood. Additionally, a year of exposure to UFSM increased height growth and nine years of school lunch exposure increase the likelihood of being of near perfect health. Exposure to school lunches also decreases the probability of being diagnosed with any health condition.

The final study by Goodchild et al. [40] investigated factors associated with a universal infant free school meal take-up and refusal in a multicultural urban community involving 676 parents of nursery aged children (4-7 years). The study explored two groups; non-provision, n = 159 or took provision, n = 517. The non-provision group were more likely to be White-British, have higher socio-economic status, have English as a first language, and involve their child in the decision over whether to take the school meal, compared to the provision group. It should be noted that the area the study was conducted is multicultural and school meals must cater for children from a variety of cultural backgrounds. Parents who did not complete questionnaires correctly were more likely to have lower socio-economic status, be non-white British and have English as an additional language, meaning some groups were under-represented.

## Discussion

A scoping review methodology in accordance with the PRISMA guidelines [29], was used to explore the global impacts of SFP on children’s health and wellbeing outcomes. To our knowledge, this is the first scoping review to comprehensively appraise SFP depending on SFP type (targeted or universal). From a health perspective, SFP positively influenced children’s physical health in terms of their social, emotional, and psychological well-being. Research on the effectiveness of FSM on BMI, weight, and nutritional intake outcomes is limited but does indicate some positive impacts. Both targeted and universal approaches play a crucial role in addressing inequalities among disadvantaged children by providing access to food; however, evidence supporting one as more effective in impacting health outcomes is limited.

In terms of physical health, most included papers investigated BMI, with some conflicting findings. For example, most papers indicated positive impacts, however two papers [6, 57] observed increased BMI under SFP. This discrepancy may be due to pre-existing malnutrition being present prior to provision or the foods served. For example, Goel et al. [55] found an increased sugar intake in free school breakfast. Though, nutritional quality of foods served differs between countries, making this difficult to assess.

SFP appears to be a positive step forward in promoting positive psychological and emotional outcomes. However, studies in this area are limited and have several limitations. It is clear that more research is needed to evaluate the effectiveness of SFP on children’s psychological and emotional outcomes.

SFP, whether universal or targeted, seems to ease food insecurity. Individuals from low socio-economic backgrounds benefited more from SFP, regardless of whether they were targeted or universal. However, targeted programs were associated with stigma, resulting in lower take-up and poorer mental health outcomes. Emphasising the importance of implementing policies to mitigate stigma. Universal systems eliminated stigma and significantly increased uptake. Nonetheless, issues such as loss of agency and food quality persisted in influencing take-up rates.

### Implications for Research

As previously mentioned, heterogeneity in research designs makes evaluating SFP difficult. Most papers included in this review were conducted over a short time span. To fully understand the impacts of SFP on children’s health outcomes, longer term research designs are essential. Additionally, sociality was deemed an important aspect of free school meals uptake, whereas stigma was associated with lower uptake. Therefore, future research should consider the implications of stigma on children’s health and how this can be alleviated. Only three studies investigated SFP as a means of addressing nutritional deficiencies among children. Future research should seek to explore these in more detail as well as other health concerns, such as respiratory illnesses and other common childhood illnesses associated with poor dietary intake.

### Implications for Practice

There is not a definitive global recommendation for SFP implementation. Based on available evidence, both targeted and universal provision appears to be effective at improving children’s health outcomes to some degree, however there are considerable limitations in current research. There is a notable need to address factors influencing uptake, particularly food quality and choice. Information was deemed important to parents when registering for SFP. Therefore, educational settings should ensure parents have sufficient information relating to SFP. Going forward, best practice and nutritional quality should be considered.

### Conclusions

Our review of 44 articles provides mixed findings on the effectiveness of SFP in improving children’s health outcomes. SFP can contribute to healthy weight outcomes, although the nutritional benefits depend on the composition of the food offered. Universal provision reduced stigma and may lead to a more positive social environment. Overall, both targeted and universal approaches were effective in addressing inequalities and food insecurity.

## Data Availability

All data relevant to the study are included in the manuscript.

## Author Contributions

Amy Locke led the review, was responsible for managing the project, analysing data and preparing an original draft.

Michaela James provided supervision, contributed to abstract and full text screening, quality assessment and editing.

Hope Jones contributed to abstract and full text screening, and quality assessment, editing. Rachel Davis contributed to abstract and full text screening and quality assessment, editing. Francesca Williams contributed to full text screening and quality assessment, editing.

Sinead Brophy provided supervision and advice for the scoping review and methodology and reviewed the final report.

