## Supplementary table 1 for "Impacts of Global School Feeding Programmes on Children’s Health and Wellbeing Outcomes: A Scoping Review"

| **Title** | **Author** | **Year** | **Country** | **Provision** | **Sample Size** | **School age** | **Outcome Measured** | **Design** | **Results** | **Limitations/Gaps** | **Recommendations** |
| --- | --- | --- | --- | --- | --- | --- | --- | --- | --- | --- | --- |
